# Young Women’s Preferences for a Self-Sampling Intervention to Diagnose Sexually Transmitted Infections: A Discrete Choice Experiment

**DOI:** 10.1101/2024.06.03.24308383

**Authors:** Ziningi N. Jaya, Witness Mapanga, Ropo Ogunsakin, Tivani P Mashamba-Thompson

## Abstract

The high rates of sexually transmitted infections (STIs) in young women in South Africa warrant the use of innovative interventions like self-sampling to diagnose both symptomatic and asymptomatic infections. Although proven as an effective measure in the fight against STIs, there is limited evidence on the preferred attributes of this intervention. We conducted a discrete choice experiment (DCE) to understand young women’s preferred attributes for self-sampling which included accessibility and convenience of self-sampling kits, education and normalisation, confidentiality and communication of results, self-sampling collection method, cost, and youth-friendliness as developed using a nominal group technique. A total of 206 young women aged between 18 – 24 years residing in underserved communities in Ethekwini Metropolitan Municipality, in KwaZulu-Natal, participated in the study. Study findings highlighted young women’s preference for enhanced accessibility, comprehensive education on STIs and self-sampling, confidential result communication, autonomy in self-collection method selection, and youth-friendly healthcare environments. The design of effective self-sampling interventions that promote STI testing thereby reducing transmission of infection, should address these preferences. Policymakers and healthcare providers should engage youth in the design of such initiatives and promote patient-centred healthcare to meet their preferences and improve STI-related health outcomes in this population.

## Introduction

South Africa faces a major public health challenge with the high prevalence of sexually transmitted infections (STIs) (1, 2, 3). With a large portion of the overall infections being among young women, early diagnosis and treatment is paramount to prevent the spread and development of the associated long term sexual and reproductive health complications (4, 5, 6, 7). Syndromic management is a long standing intervention used to provide STI healthcare services, particularly in low-and-middle-income countries (LMICs), like South Africa, where quality healthcare is not easily accessible to all (8, 9). Although widely used, this intervention is unable to detect asymptomatic infections which are common with STIs. By requiring patients to visit a healthcare facility for medical assistance, it deters patients for various reasons (10). Some common deterrents include the discomfort with associated invasive genital examinations and fear of judgement for being sexually active and the stigma associated with STIs (10, 11). Furthermore, since syndromic management does not rely on laboratory confirmation to administer treatment, it often leads to over-diagnosis and over-treatment of STIs which may promote drug resistance. Antimicrobial resistance is a growing global concern (12, 13), and STIs are not exempt. Several researchers have reported on the growing trend of drug resistant gonorrhoea and chlamydia (14, 15, 16) which are both curable bacterial STIs. Considering this there is an urgent need for innovative accessible interventions to promote and improve healthcare seeking behavior and minimise the development and spread of antimicrobial resistande, especially among young women.

Self-sampling for STI diagnosis as an intervention that eliminates the main challenges presented by syndromic management, has gained recognition. Self-sampling interventions have been proposed as a potential solution to eliminate challenges presented by syndromic management and increase access to STI screening services for young women in underserved communities (17, 18). Although not used in practice in South Africa, and most LMICs, its ability to eliminate the main challenges posed by syndromic management is well understood. Furthermore, it does not only increase healthcare access in resource-limited settings but also enables screening and testing of asymptomatic infections (19, 20). When considering the reported STI crisis among young women and the challenges that negatively impact their healthcare seeking behaviour, it is clear that an intervention that is pacceptable and accessible would prove useful. Self-sampling as an intervention, appears to be a good alternative to enable easy access and promote STI testing within this population. Based on this, a model for providing this intervention according to user preferences, particularly in the South African context, would prove beneficial.

Choice experiments have been utilised to understand decision-making processes and people’s preferences in various contexts. A common example of such experiments is the attribute-centred discrete choice experiment (DCE) in which individuals’ trade-offs between attributes are quantified to determine user preferences (21). The ultimate purpose of these experiments is to reveal the extent to which an individual is willing to give up to benefit more from another attribute (22, 23, 24). Considering the need for an STI healthcare model that is acceptable for young women, a DCE is the ideal approach to ascertain young women’s preferences for a self-sampling intervention for STI diagnosis. Therefore, a DCE was conducted among young women aged 18 – 24 years, to establish their preferences for a self-sampling intervention to diagnose STIs. We investigated trade-offs between attributes which included accessibility and convenience of self-sampling kits, education and normalisation, confidentiality and communication of results, self-sampling collection method, cost, and youth-friendliness as developed using a nominal group technique.

The young women were selected from underserved communities in eThekwini Metropolitan Municipality, an area with the highest population density in KwaZulu-Natal. In South Africa, the province of KwaZulu-Natal constitutes the largest portion of people with STIs (25). Based on this, it was an ideal site for the study to be conducted. Understanding young women’s preferences can inform policy and guide the development of more effective user-friendly interventions to address the existing STI burden. This could reduce stigma, increase STI testing uptake, improve healthcare outcomes, and ultimately align with Sustainable Development Goal (SDG) 3 to ensure universal access to sexual and reproductive healthcare services (26).

## Methods and analysis

### Study design

A DCE is a quantitative data collection method employed to determine user or participant preferences for a service or goods provided. The DCE is founded on three theories including the random utility theory (RUT), Lancaster’s characteristics theory of demand, and the standard microeconomic theory of consumer (27). The random utility theory suggests that there is a “true” utility that individuals derive from consuming products, but this utility cannot be fully observed by researchers (28). Instead, it is summarized by two components: a systematic (explainable) component and a random (unexplainable) component (28). The Lancaster’s characteristics theory on the other hand suggests that product utility doesn’t lie in the actual product instead it lies in the characteristics of the product (29). According to the standard microeconomic theory the individual sensibly seeks to maximise utility using available information and constraints (30). The DCE process determines preference based on a series of attributes with a series of associated options or choice tasks for users to select (23, 31, 32). In this way, researchers can ascertain the level of importance that participants place on each attribute which illuminates the trade-offs they are willing to make on one attribute versus another (22, 23).

### Selection of attributes and levels

A DCE survey was conducted on young women residing in underserved communities in eThekwini Metropolitan Municipality between February to March 2024. In the DCE, participants completed a survey comprising a series of *choice tasks* presented with hypothetical scenarios for attributes of a self-sampling intervention for diagnosing STIs. The attributes consisted of different levels. Participants were presented with 2 different alternatives of attribute configurations to choose from for each scenario. Prior to conducting the survey, attributes were co-created with healthcare workers from primary healthcare clinics (PHCs) and young women in underserved communities using a nominal group technique (NGT) (33, 34). After the NGTs, a total of eight attributes emerged namely accessibility, education, communication, convenience, youth-friendliness, and cost of the self-sampling kit. We opted to utilise the vaginal swab and urine collection as the self-sample collection methods. This decision was informed by literature which highlights these collection methods as non-invasive (35) and the most commonly used for STI diagnosis in women (36, 37). After an expert research panel review of all the attributes, a few of these were merged resulting in six attributes for use in the survey. The final attributes included accessibility and convenience, education and normalisation, confidentiality and communication, self-sampling method, youth-friendliness, and cost of self-sampling kits. See Table 1 for attribute description and their levels.

**Table 1:** Attribute description and levels.

| Attribute<br>(regression<br>label) | Description | Levels ( or preference parameters) |
| --- | --- | --- |
| **Accessibility**<br>**and**<br>**Convenience**<br>**(access)** | Refers to the ease with which young women can obtain self-sampling kits for STI screening and the level of convenience in the process. | • Kits are only available at clinics.<br>• Kits are available at clinics, schools, and pharmacies.<br>• Kits are available through outreach, clinics, schools, pharmacies, and with online support. |
| **Education and**<br>**normalisation**<br>**(education)** | Refers to the level of information and awareness provided to young women about STIs and self-sampling, as well as education efforts to reduce stigma and promote testing. | • No educational material on STIs is provided with the kit.<br>• Basic information on STIs is provided with the kit.<br>• Comprehensive information is provided with kits and campaigns in the community to encourage testing. |
| **Confidentiality**<br>**and**<br>**communication**<br>**(confid)** | Focuses on how screening and testing results are handled, focusing on the level of privacy and mode of result communication. | • Results are given at the clinic.<br>• Remote sharing of results via phone, text, email, or online portal. |
| **Self-sampling**<br>**collection**<br>**method (self)** | Refers to the sampling kit or tool used to collect the specimen. | • Vaginal swab kit issued.<br>• A urine sample kit is issued.<br>• Kit offers a choice of methods (swab or urine). |
| **Youth-**<br>**friendliness**<br>**(youth)** | Improving youth-friendly services at clinics could help to make the experience more comfortable and welcoming for young women. | • No improvements were made to create a youth friendly environment for young people during clinic visits.<br>• Clinics make minor improvements to create a youth friendly environment (eg. separate areas, comfortable rooms, and trained staff). |
|  |  | • Clinics make major improvements and efforts to create a youth friendly environment (eg. extended hours, dedicated youth-friendly space). |
| **Cost of self-sampling kits (cost)** | Making the self-sampling kits available free of charge at local pharmacies, mobile clinics, schools, and universities could remove financial barriers to accessing STI screening services. | • Kits need to be purchased.<br>• Kits are free at clinics only.<br>• Kits are free at clinics, schools, pharmacies, and mobile clinics. |

### Questionnaire design

#### Pilot study

A total of 20 participants were involved in the pilot study to pre-test the survey tool and determine the robustness and feasibility of the DCE. The pilot study was used to determine whether the attributes and levels were relevant, and understandable to participants, and the overall clarity of the questionnaire. It also sought to determine the amount of time it took to complete the tool in order to determine the level of burden that participation would place on those who agreed to participate. The overall aim was to make it as simple and convenient for the participants. Participants reported ease and no comprehension challenges. A majority of the participants (80%) reported the length of the tool as a challenge and suggested a reduction in the number of choice tasks. Participants agreed on the relevance of suggested attributes and made no suggestions for additional ones. The tool was amended accordingly based on participant comments. Thereafter it was reviewed once again by 5 volunteers, who met the participant eligibility criteria, and deemed it suitable for data collection.

#### Experimental design

The DCE had six attributes (5x3 levels, 1x2 levels), meaning that there were (3 x 3 x 3 x 3 x 3 x 2) = 486 different potential combinations of attributes, *choice tasks*, and levels in a full factorial design. However, this number of choice tasks is too high and would prove burdensome for the participants. Based on this, we utilised an efficient design which includes both random parameters and an error component in the utility functions using Ngene software. This design choice was particularly suited to our sample size of 196, and aimed to reduce participant burden while maintaining statistical rigor. Random parameters capture individual-level variability in preferences, while error components account for unobserved sources of variability and stochasticity in choices. The experimental design yielded 12 choice tasks which offered participants two options to choose from. Since there is no prior evidence of such an investigation, having two options in the survey was suitable to gain a nuanced understanding of young women’s preferences and perceptions of a self-sampling intervention to diagnose STIs. It further enabled the collection of foundational insights into the subject of investigation.

Additionally, the simplicity of this approach reduces the cognitive load on the participants leading to effortless decision making with potentially more reliable responses. In addition to the 12 experimental choice tasks, two additional choice tasks were added. One of these choice tasks was for the participants to practice before completing the actual survey and the other was a repeat of the third choice task to assess consistency (38). Therefore, participants completed 14 choice tasks.

#### Questionnaire design

The questionnaire comprised three sections. Section one was mainly to collect demographic information about the participants including age, knowledge about STIs, and socioeconomic information. Section two provided introductory guidance and examples on how to complete the survey including a practice choice task. They were asked to imagine the following scenario as a way to stimulate their thoughts about the choice they would make to become familiar with the process for completing the survey:

*You are a young woman who recently had unprotected sex with a man you do not trust. Concerned about the potential risk of STIs, you find yourself contemplating getting tested. The decision weighs heavily on your mind as you consider various factors that might influence your choice*.

Each of the participants then proceeded to complete all 14 choice tasks. Following the completion of the choice tasks, section three presented statements describing and enquiring about the participants’ experience with understanding the DCE survey in terms of relevance, significance, clarity and complexity.

### Participants

Participant selection and eligibility were based on participants being aged between 18 – 24 years, residing in an underserved community in the selected study areas, and having knowledge about STIs. All interested participants provided written informed consent before the completion of the survey. Those who were not willing to provide consent were excluded from participating in the study. All consenting participants answered screening questions to determine eligibility before completing the survey. Thereafter, a walkthrough of the survey questions was provided to facilitate understanding and ease of completion. Due to safety concerns, a paper based survey was completed and the information gathered was later transferred onto an Excel spreadsheet. However, where possible the survey was administered electronically using Google form. Data collection using the DCE surveys was conducted from 01 February to 06 March 2024.

### Data analysis

The demographic and socioeconomic characteristics of the respondents are described. Categorical data is presented by absolute and relative frequencies (n and %). Bivariate logistic regression was conducted to model participant preferences for self-sampling attributes. Binary outcome variables indicating preference or non-preference for each attribute level were used with predictor variables. The regression coefficients, odds ratios, and statistical significance of predictor variables were analyzed to determine the preferences of participants for different self-sampling attributes. Additionally, the kappa coefficient was calculated to measure agreement regarding ease of understanding of the survey tool. Model fit was assessed using both a log-likelihood ratio (LLR) test, Aikaike’s Information Criterion (AIC), and Bayesian Information Criterion (BIC) (39). Model selection criteria, such as the Akaike Information Criterion (AIC) and Bayesian Information Criterion (BIC) were reported.

A DCE protocol was developed for this study and is accessible on https://www.medrxiv.org/content/10.1101/2024.01.05.23299719v1

## Results

### Participant characteristics

The target sample size for the DCE was 196, however, a total of 206 young women aged between 18 – 24 years participated in the survey. This means study participation was 105%. In this research, the demographic characteristics, socioeconomic status, and healthcare knowledge of participants were studied. Most participants were aged 18-21 years (53%) and 22-24 years (47%). Household income varied, with notable percentages in the R1-R4800/month (40%) and R4801-R9600/month (38%) brackets. Urban formal settlements were more common (69%) than urban informal ones (31%), and a majority of participants owned their dwellings (70%). High levels of STI awareness (98%) and knowledge about local healthcare services (99%) were observed. However, a significant portion of participants (32%) expressed discomfort with available healthcare services, suggesting areas for improvement in service delivery or patient experience. **Table 2** below presents a summary of participant characteristics.

**Table 2:** Summary of demographic, socioeconomic status, and healthcare knowledge.

| Variable name | Categories | Frequency | Percentiles |
| --- | --- | --- | --- |
| Age | 18-21 years | 109 | 53 |
|  | 22-24 years | 97 | 47 |
| Household Income | No Income | 32 | 16 |
|  | R1 - R4800/Month | 81 | 40 |
|  | R4801 – R9600/Month | 78 | 38 |
|  | R9601 – R19200/Month | 12 | 5 |
|  | >R19200/Month | 3 | 1 |
| Dwelling Type | Urban Formal Settlements | 142 | 69 |
|  | Urban Informal Settlements | 64 | 31 |
| Dwelling Status | Owned | 145 | 70 |
|  | Rented | 61 | 30 |
| STI Awareness | Yes | 201 | 98 |
|  | No | 5 | 2 |
| Knowledge about local STI healthcare services | Yes | 204 | 99 |
|  | No | 2 | 1 |
| Feelings about the available healthcare service | Comfortable | 140 | 68 |
|  | Uncomfortable | 66 | 32 |

### Young women’s preferences for a self-sampling intervention

Figure 1 reports the model fitness and participant preferences for the self-sampling attributes. Model fit was assessed using both a log-likelihood ratio (LLR) test, Aikaike’s Information Criterion (AIC), and Bayesian Information Criterion (BIC). The logistic regression model exhibited robust fit as indicated by the log-likelihood (-3712.87) and information criteria (AIC: 6377.74, BIC: 6456.77). For accessibility and convenience, participants were more likely (OR 2.892 p<0.001) to opt for kits to be available through outreach, clinics, schools, pharmacies, and online support compared to clinics only. For education and normalisation, participants were more likely to opt for comprehensive information on STIs and self-sampling to be provided with the kits and community campaigns compared to not having any information at all. Concerning confidentiality and communication, the participants were 1.696 (p<0.01) more likely to opt for the sharing of results via text, phone, email, or using an online platform compared to collecting results from the clinic. For the self-sampling collection method, our participants were 1.441 (p<0.01) more likely to opt for the urine self collection method compared to the vaginal swab.

**Fig 1:**
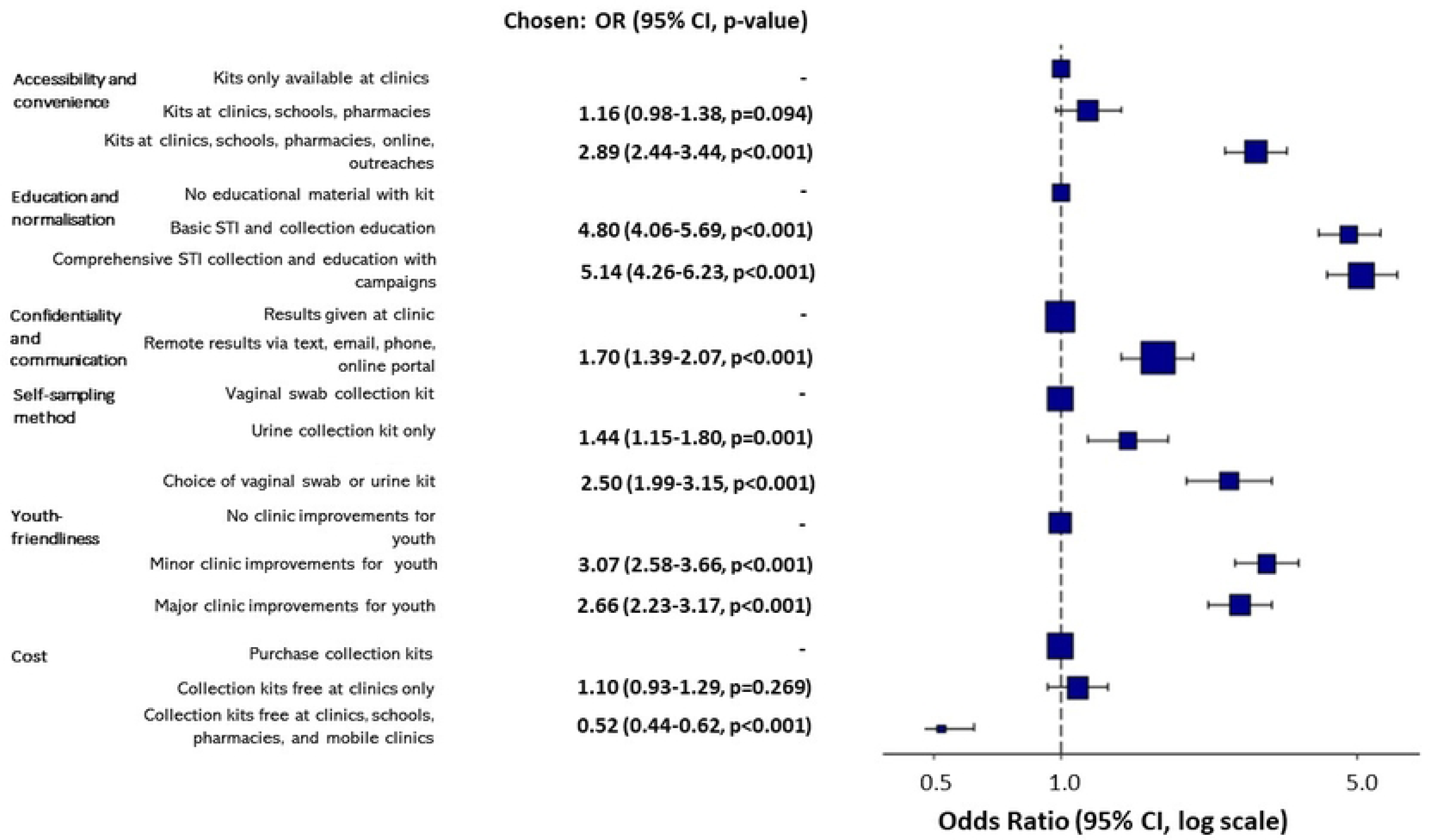
Summary of preferences and model fitness. Note: Significance: *** P-value < 0.001, ** P-value < .01, * P-value < .05; OR: Odds ratio

Additionally, for the same attribute, another portion of participants were more likely (2.659 p<0.001) to prefer the option to choose between urine or vaginal swab self-collection compared to only self-collecting a vaginal swab. Concerning the cost of self-sampling kits, 48% (OR 0.523 P<0.001) of the participants were less likely to opt for free kits at clinics, schools, pharmacies, and mobile clinics compared to kits being purchased.

### Clarity and robustness of the survey

Additional variables were evaluated to ascertain participant experiences with the survey tool to determine the clarity and robustness of the survey, see Fig 2 below for a summary. Regarding the understanding of the survey, 202 participants (98%) agreed that they found the survey easy to understand, while 3 participants (1.5%) were uncertain, and only 1 participant (0.5%) disagreed.

**Fig 2:**
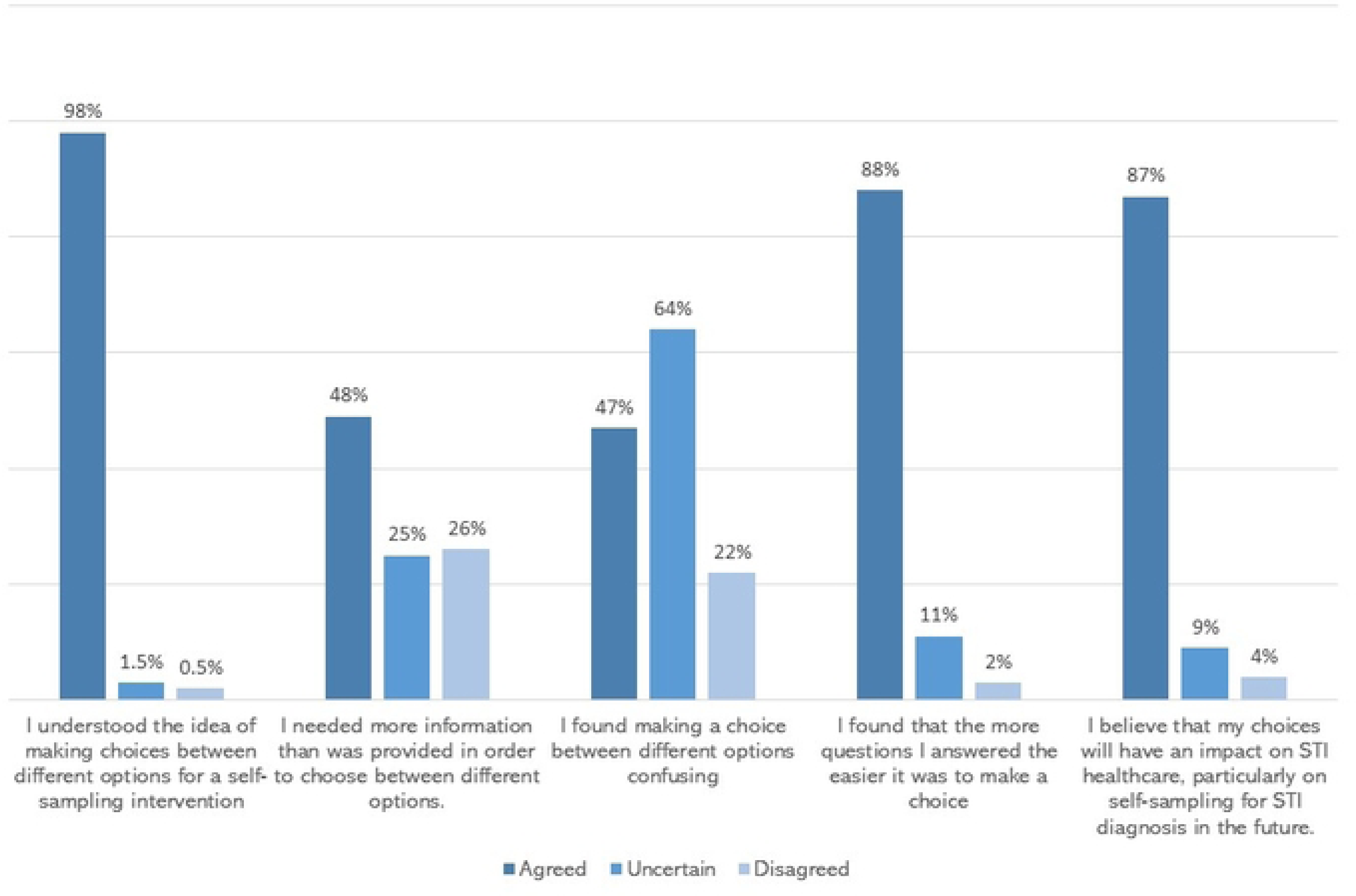
Summary of survey tool clarity and robustness.

In terms of information needed to inform decision-making between choice set options, 102 participants (49%) agreed that they needed more information, 51 participants (25%) were uncertain, and 54 participants (26%) disagreed with the need for additional information.

When considering confusion between competing options, 97 participants (47%) did not find it confusing, 64 participants (31%) were uncertain, and 45 participants (22%) agreed that they were confused when deciding between options. Regarding the perceived ease of answering more choice sets, 181 participants (88%) agreed that answering more questions improved their ability to make a choice, while 24 participants (11.5%) were uncertain, and 1 participant (0.5%) disagreed with this notion. Only 180 (87%) participants believed their study participation would impact the provision of STI healthcare services, 18 (9%) were uncertain, and only 8 (4%) did not believe so.

Using this data, the kappa coefficient was calculated to assess the level of agreement beyond chance among participants across these variables. The kappa coefficient was approximately 0.456, indicating moderate agreement beyond what would be expected only by chance.

## Discussion

The main objective of this study was to conduct a DCE among young women in underserved communities to understand their preferences for a self-sampling intervention to diagnose STIs. Most of the young women who participated in our study were aged under 21 years. This indicates that there is a large portion of youth with specific healthcare needs and preferences in relation to STIs. Considering that STI prevalence is reportedly high among young people across the globe (40, 41), the proportion of young women who participated is of great significance. It is important to note that some of the participants had no income or were from lower middle-income households, having little to no household income. This is not alarming considering the World Bank report which reports on a high concentration of poverty in Sub-Saharan Africa (42). When reviewed in conjunction with the notable portion of participants from rented urban informal settlements, the influence of these socioeconomic nuances on STI prevalence and accessibility of healthcare is clear. Living in poverty may lead to reckless sexual behaviour stemming from desperate circumstances for survival (43, 44), where reckless sexual behaviour increases the risk of STI acquisition.

Our study highlights a preference for self-collection kits to be widely available through various channels, including outreach programs, clinics, schools, pharmacies, and online platforms to enhance access and convenience of self-sampling interventions. This finding underscores the significance of having a multi-faceted approach to accessibility to such STI screening interventions to promote uptake and utilization among young women. This aligns well with previous studies that demonstrate increased self-care which increases uptake when healthcare services are easily accessible (45). The more convenience and ease of access to a service, the higher the chances of use or engagement with it. There was a strong preference for comprehensive educational materials to accompany the self-collection kits highlighting the role of education in normalizing STI-related screening and healthcare seeking behaviour. This aligns with existing literature which supports educating the public on sensitive issues, such as those related to sexual health, to reduce STIs and empower individuals to make better informed decisions (46, 47). As such, the provision of detailed information about STIs and self-sampling procedures not only empowers individuals to make informed decisions but also contributes to reducing stigma and further increases awareness about sexual health in this population age group.

The importance of maintaining confidentiality and privacy of STI diagnosis for the chosen population was demonstrated by their preference to receive diagnostic test results through modern communication channels such as text messages, emails, phone calls, or online portals. This mode of result communication not only maintains confidentiality but also ensures timely access to results which leads to timely access to treatment. Furthermore, this reduces interaction with healthcare providers thereby addressing previously reported fear of judgement and stigma by healthcare providers as a barrier to seeking medical care for STIs. Concerning the self-collection method, our findings suggest participants have a strong desire for autonomy and choice in the self-sampling method. Having the option to choose between different collection methods, instead of being restricted to a single method, empowers individuals to make choices that suit their preferences. This resonates with the principle of patient-centred care that encourages healthcare provision in partnership with patients recognising their preferences and values (48).

The preference for designated youth-friendly areas at healthcare facilities highlights the importance of creating an inclusive environment suitable for youth. For instance, having a space dedicated to youth, staff trained to deal with young people, comfortable rooms, and extended clinic hours can potentially reduce reluctance and increase utilization of STI-related medical services. This aligns with studies that highlight the need for designated youth-friendly services to improve the delivery of STI related services to young people (49). Interestingly, our study highlighted that the option to purchase or receive kits for free had no significant influence on participant preferences when compared to the other attributes. This suggests that while cost may be an important factor to consider, the other attributes may play a more significant role in shaping individual preferences and behaviours related to STI screening and diagnosis, for self-sampling.

### Policy, practice, and research implications

The study findings have important implications for policy, practice and further investigation in the area of self-sampling interventions for STI diagnosis in young women. Concerning policy, there is a clear need to increase focus on initiatives that enhance the accessibility of self-sampling kits using various approaches including clinics, schools, community outreach programmes, and online platforms. Policies should also advocate for the development and implementation of comprehensive education initiatives to provide clear and accurate information about STIs and self-sampling procedures. This would help to promote testing for STIs, even among asymptomatic individuals and reduce associated stigma. Policy frameworks should also emphasise the importance of maintaining confidentiality and privacy regarding the communication of diagnostics results using secure channels including text, phone calls, and online platforms where possible. Additionally, policy should advocate for the creation of youth-friendly environments in healthcare facilities to enhance engagement with STI-related healthcare services by young people.

In relation to practice, a patient-centred approach should be adopted by offering options and autonomy in self-sampling collection methods. Individuals engaging in self-sampling interventions should be provided with comprehensive education and counselling to address concerns that arise and also cultivate a culture of regular testing to improve healthcare seeking behaviour. Additionally, to ensure that services are utilised it is essential to engage young people in the design and implementation of self-sampling for screening and diagnosis of STIs.

We recommend a longitudinal study to assess the long-term impact of preferred self-sampling intervention attributes on the uptake of such an intervention for STI diagnosis and health outcomes among young women. A comparative effectiveness study to evaluate different models of the intervention is also recommended. Future research should consider health equity and technology integration ineffectively delivering self-sampling interventions to diverse communities. Ultimately, there is a great need for policy and practice interventions that address young women’s preferences and needs concerning self-sampling to improve STI-related healthcare seeking behaviour and reduce rates of STI transmission.

## Limitations

Our study findings may be limited in their application to the broader community of young women because we did not investigate certain demographic information including employment status, source of income, marital status, and parental status. Data was only collected among young women residing in underserved urban communities, who may have unique circumstances that do not necessarily apply to those residing in other types of communities. As such, our findings may only apply to young women residing in other areas similar to those of the study. Our investigation only focused on a set of attributes and there is a potential that other factors and preferences that could influence participant behaviour and decision-making for STI diagnosis were overlooked.

## Conclusion

In conclusion, the study demonstrates a clear preference for enhanced accessibility, comprehensive education on STIs and self-sampling, confidentiality in results communication, individual autonomy in self-collection method selection, and youth-friendly environments in healthcare facilities. These preferences underscore the need to address key attributes to design self-sampling interventions that effectively promote STI screening and diagnosis behaviour which ultimately reduces rates of transmission. As a result, there is a need for policymakers and healthcare providers to engage young people in designing and delivering STI-related services, prioritize initiatives to improve accessibility, support education on STIs and self-sampling education, ensure confidentiality and privacy in result communication, create youth-friendly environments, and promote patient-centred care.

## Data Availability

All relevant data are within the manuscript and its Supporting Information files.

## Acknowledgements

We thank participants for their time and commitment to completing the survey.

## Author contributions

**Conceptualization:** Ziningi Nobuhle Jaya, Tivani Phosa Mashamba-Thompson.

**Data curation:** Ziningi Nobuhle Jaya.

**Formal analysis:** Ziningi Nobuhle Jaya.

**Funding acquisition:** Ziningi Nobuhle Jaya.

**Investigation:** Ziningi Nobuhle Jaya.

**Methodology:** Ziningi Nobuhle Jaya, Tivani Phosa Mashamba-Thompson.

**Project administration:** Ziningi Nobuhle Jaya.

**Resources:** Ziningi Nobuhle Jaya.

**Software:** Ropo Ogunsakin and Ziningi Nobuhle Jaya.

**Supervision:** Tivani Phosa Mashamba-Thompson, Witness Mapanga.

**Validation:** Ziningi Nobuhle Jaya, Tivani Phosa Mashamba-Thompson.

**Visualization:** Ziningi Nobuhle Jaya.

**Writing – original draft:** Ziningi Nobuhle Jaya.

**Writing – review & editing:** Tivani Phosa Mashamba-Thompson, Witness Mapanga.

## Acknowledgements

We would like to thank the healthcare workers and young women who participated in the DCE and those who assisted with data collection.

## Authors’ contributions

Conceptualization, Z.N.J. and T.M.-T.; data collection; writing - original draft, Z.N.J.; writing—reviewing and editing, T.M.-T., and W.M; Statistical analysis: R.O and Z.N.J, supervision, T.M.-T and W.M.

## Patient consent for publication

obtained.

## Ethics approval

Ethical clearance was obtained from the University of Pretoria Research Ethics Committee (reference number 136:2022) and the KwaZulu-Natal Department of Health (reference number KZ_202208_005) prior to data collection. All participants signed informed consent prior to data collection.

## Data availability statement

no additional data is available.

